# Silent Battles: The Impact of War on the Mental and Physical Health of Endometriosis Patients

**DOI:** 10.1101/2025.03.07.25323565

**Authors:** Ohad Regev, Sharon Livneh, Noam Schuman Harel, Amit Eliyahu, Beatris Pekar Agronsky, Shir Shahar, Aya Wertheimer, Dana Lassri

## Abstract

**Background:** Endometriosis is a chronic, inflammatory disease affecting 1 in 10 women of reproductive age worldwide. It is characterized by a range of debilitating symptoms which collectively impair patients’ quality of life. While stress is a well-documented factor known to exacerbate endometriosis symptoms, the impact of extreme and prolonged external stressors, such as ongoing war, on disease progression and patient well-being hasn’t been thoroughly studied.

On October 7, 2023, Israel faced a severe terror attack impacting Jewish and non-Jewish victims alike. Since then, Israel has been involved in an on-going conflict on multiple fronts. This study explores the compounded effects of ongoing war on women with endometriosis, focusing on symptom severity, physical and mental health, and illness perception.

**Materials & Methods:** A cross-sectional survey study was conducted in Israel in August 2024, recruiting 841 women with confirmed diagnosis of endometriosis and/or adenomyosis through social media platforms and endometriosis clinics. The survey assessed the impact of the war on participants’ lives, physical, and mental health. It consisted of several validated measures as well as sociodemographic and clinical questions. Statistical analyses included descriptive statistics, univariate statistics for assessing temporal changes in symptoms severity and perceptions across three timepoints (pre-war, 2 months post-war, ∼1 year post-war), and correlation between war-related stressors, mental status, and health outcomes. Univariate and multivariable logistic regression models were employed to identify factors associated with symptom worsening, adjusting for sociodemographic characteristics, medical conditions, and war-related factors.

**Results:** 82.8% of participants were directly affected by war-related stressors. Mental health deteriorated substantially, with anxiety rates increasing from 34.7% to 94.1% and depression from 39.6% to 89.3% (p<0.001). Physical health was also affected, with 77.4% reporting worsening endometriosis symptoms and 62.5% indicating overall medical decline. All specific symptoms, including pelvic pain, digestive and urinary symptoms, dyspareunia, fatigue and more, showed significant deterioration (p<0.001). Additionally, 38.9% and 14.4% of participants reported increased usage of pain and hormonal medications, respectively, indicating higher symptom-management needs. Multivariable analysis revealed significant associations between worsening in symptom severity to war-related stressors (aOR=1.24, 95%CI=1.07-1.45), war-related stress levels (aOR=1.83, 95%CI=1.50-2.22), depression and anxiety levels (aOR=2.00, 95%CI=1.52-2.63), and impaired healthcare accessibility (aOR=1.64, 95%CI=1.04-2.57). Negative illness personification was associated with worse outcomes (aOR=1.37, 95%CI=1.12-1.67), while positive illness personification showed protective effects (aOR=0.77, 95%CI=0.59-0.99).

**Conclusions:** This study demonstrates the profound impact of war and war-related stress on both the physical and mental health of endometriosis patients, highlighting the critical need for targeted healthcare interventions and psychological support during times of conflict.

## Background

Endometriosis is a chronic and often debilitating disease affecting millions of women globally. It is characterized by the growth of endometrial-like tissue outside the uterus, leading to persistent pelvic pain, irregular menstrual cycles, dysmenorrhea, dyspareunia, infertility, and chronic fatigue.^1–3^ Beyond its effect on patients’ physical health, there is a growing recognition of the profound impact of the disease on patients’ mental health and psycho-social well-being. The persistent and often severe nature of the symptoms, along with the long diagnostic delay, have been shown to effect patients’ quality of life, mental health, self-esteem, relationships, and work productivity.^4–12^

Recent studies suggest that stress, and especially chronic stress, can exacerbate endometriosis symptoms.^13–18^ Moreover, research in animal models revealed that stress can influence the hormonal balance and immune response, leading to increase in the size and severity of the endometrial lesions and exacerbation of inflammatory parameters.^19,20^ Also, stress was found to promote nerve fiber growth in the uterus, suggesting that increased density of nerve fibers may play an important role in the pathogenesis of pain and tenderness.^21,22^ Conversely, stress reduction has been shown to decrease lesion size by nearly 50%.^22,23^

War-related stress has been shown to have profound psychological impact, contributing to elevated rates of depression, anxiety, and post-traumatic stress disorder (PTSD), even among individuals indirectly exposed to traumatic events through media.^24–29^ On October 7, 2023, Israel experienced an unprecedented attack by Hamas involving extensive rocket assaults and large-scale terrorist infiltration into southern Israel, leading to over 1,200 casualties and the abduction of 251 civilians of all ages and ethnic backgrounds.^30,31^ This attack, the deadliest act of terrorism in Israel’s history,^32^ was marked by severe acts of violence, including torture, maiming, and rape, many of which were broadcast live on social media by the terrorists, amplifying psychological trauma on a national scale.^33–35^ Since the initial assault, Israel has engaged in ongoing war with several militant groups, including Hamas, Hezbollah, the Houthis, and Iran. The conflict has led to mass civilian evacuations, extensive casualties, and profound national trauma, with recent studies highlighting increased rates of PTSD, anxiety, depression, distress, and substance abuse.^36–39^ Moreover the ongoing conflict disrupted healthcare services in Israel, with medical centers closed, healthcare providers called to military service, and patients displaced from their homes. The interruption in healthcare services risks substantial delays in diagnosis and treatment for endometriosis, potentially worsening physical symptoms and exacerbating psychological distress in this vulnerable group.

While studies suggest that stress may play a significant role in endometriosis exacerbation, little is known about the effect of environments marked by extreme external stressors, such as war and conflict on the disease. To date, only one study from Israel explored this topic; however, it was limited by a small sample size and was conducted in a single center, reducing its generalizability to the broader population of women with endometriosis.^40^

This large population-based cross-sectional study aims to explore the impact of war-related psychological stress, healthcare delays, and mental health challenges on women with endometriosis during the ongoing conflict in Israel. By examining the interplay between external stressors, healthcare disruptions, and disease progression, this research aims to provide essential insights into how national crises affect the physical and mental health of individuals with chronic conditions like endometriosis.

## Methods

### Study Design & Population

This cross-sectional survey study was conducted in Israel in August 2024. The study population consisted of women aged 18 years and older with a confirmed diagnosis of endometriosis and/or adenomyosis from a qualified medical provider.

Participants were recruited through social media platforms (dedicated endometriosis communities on *Facebook, Instagram*, and *LinkedIn*), dedicated *WhatsApp* support groups, and designated endometriosis clinics throughout Israel. The survey was administered through *Qualtrics*, an encrypted research platform that ensured data security and participant privacy. Prior to survey access, participants reviewed comprehensive study information and provided electronic informed consent.

### Measures

The survey assessed the impact of the war on participants’ lives, physical health, and mental health. It consisted of several validated measures as well as sociodemographic and clinical questions.

#### Physical Health

1. *Participants’ medical condition status*: assessed by the general self-rated health (GSRH), a single-item health measure with five options between “very good” and “very bad.” This measure is wildly used in research and has been shown to be consistent with patients’ objective health status.^41^
2. *Endometriosis symptoms severity*: Participants stated whether they had symptom deterioration in general since the beginning of the war. In addition, they rated the severity of various symptoms at three timepoints (pre-war, 2 months post-war, ∼1 year post-war). Symptoms severity was assessed on a 5-point scale, with 1 being the best and 5 being the worst.

#### Mental Health

1. *Anxiety & depression*: assessed by the Patient Health Questionnaire-4 (PHQ-4), a validated ultra-brief screening tool for anxiety and depression. The total score is determined by adding together the scores of each of the 4 items. Scores are rated as normal (0-2), mild (3-5), moderate (6-8), and severe (9-12). Total score ≥3 for first 2 questions suggests anxiety. Total score ≥3 for the last 2 questions suggests depression.^42,43^
2. *Illness personification*: Relying on anthropomorphism research, Illness Personification Theory (ILL-PERF) posits that individuals living with a chronic illness ascribe human-like characteristics to their illness. We assessed illness personification using the Ben-Gurion University Illness Personification Scale (BGU-IPS) which is a 12-item measure of negative and positive illness personification (6 items each). Each item is assessed by a 5-point scale with 1 being “Highly Disagree” and 5 being “Highly Agree”. Each personification dimension (e.g., negative IPS and positive IPS) is scored as the average of the six items. Negative personification is associated with pain intensity and illness-related distress (e.g., depression and low adjustment to pain). Positive personification is correlated with hope, pain-related sense of control, and low depression.^44–46^
3. *War-related stress:* War-related stress was measured using a 4-item scale focusing on subjective threat perception.^47^ Participants were asked to rate on a 5-point Likert scale (0 = not at all, 4 = very much) how much they felt since the war outbreak that: “your life was in danger”; “the lives of their family members were in danger”; “your family members were in danger of being hurt/injured”; “I felt helpless”. The internal consistency coefficient for this scale was strong (α Cronbach’s = 0.89). The total score is determined by adding together the scores of each of the 4 items. Scores are rated as normal (0-2), mild (3-5), moderate (6-8), severe (9-12), and very severe (≥13).

#### War Impact

The impact of the war on participants was assessed through dichotomous questions (yes/no) assessing exposure to war-related events, changes in daily activities and routines, and healthcare access disruptions (**Supplementary Tables 1+2**).^47^

### Statistical Analysis

Descriptive statistics were employed to analyze the attributes of the study population. Each variable is presented by the most suitable central and dispersion measures: dichotomous and nominal variables are presented by number and percentages and numerical variables by either mean ± standard deviation or median and interquartile range (IQR). We conducted univariate analysis using non-parametric Friedman tests and chi square tests for assessing temporal changes in symptoms severity, anxiety & depression levels, and illness perceptions across three timepoints (pre-war, 2 months post-war, ∼1 year post-war). Spearman test was used to examine the correlation between the number of war-related stressors to participants’ anxiety & depression levels, their perceptions of their disease, and their medical condition. Next, we used univariate logistic regression to assess the association between participants’ characteristics to worsening in endometriosis symptoms severity and medical condition following the war (**Supplementary Tables 3+4**). Finally, we used multivariable logistic regression to examine the impact of war-related stressors, participants’ mental health statute, and impaired healthcare accessibility on participants’ physical health after adjusting to potential confounders detected in the univariate analysis.

All statistical analyses were conducted using SPSS Statistics version 25 and R software. A two-sided test significance level of 0.05 was used throughout the entire study.

### Ethics Approval

The study was approved by the ethic committee of the School of Social Work and Social Welfare at the Hebrew University of Jerusalem per the Declaration of Helsinki.

## Results

Overall, a total of 841 women diagnosed with endometriosis completed the survey (**Table 1**). The mean age was 31.4±8.1 years. Participants were predominantly Jewish (96%), secular (65%), married or in a relationship (62.9%), and with higher education (51.3%). The cohort exhibited diverse clinical characteristics, with varying types of endometrioses. The most prevalent subtypes were superficial endometriosis (40.5%) and adenomyosis (39.1%), followed by deep endometriosis (30.9%). 43.8% had additional chronic comorbidity.

**Table 1.**
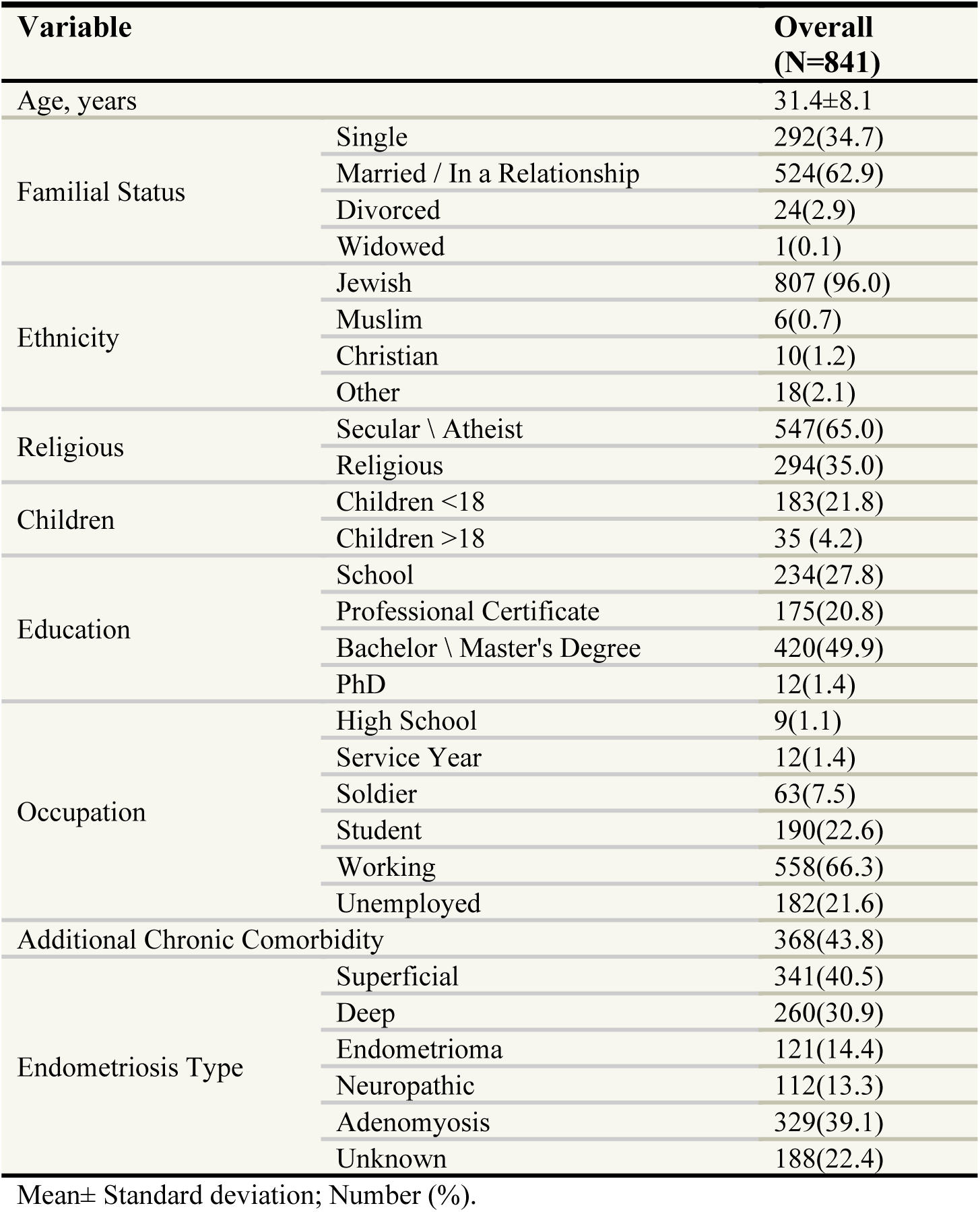
Sociodemographic & Clinical Characteristics of Study Cohort.

| **Table 1. Sociodemographic & Clinical Characteristics of Study Cohort** |  |  |
| --- | --- | --- |
| **Variable** |  | **Overall**<br>**(N=841)** |
| Age, years |  | 31.4±8.1 |
| Familial Status | Single | 292(34.7) |
|  | Married / In a Relationship | 524(62.9) |
|  | Divorced | 24(2.9) |
|  | Widowed | 1(0.1) |
| Ethnicity | Jewish | 807 (96.0) |
|  | Muslim | 6(0.7) |
|  | Christian | 10(1.2) |
|  | Other | 18(2.1) |
| Religious | Secular \ Atheist | 547(65.0) |
|  | Religious | 294(35.0) |
| Children | Children <18 | 183(21.8) |
|  | Children >18 | 35 (4.2) |
| Education | School | 234(27.8) |
|  | Professional Certificate | 175(20.8) |
|  | Bachelor \ Master's Degree | 420(49.9) |
|  | PhD | 12(1.4) |
| Occupation | High School | 9(1.1) |
|  | Service Year | 12(1.4) |
|  | Soldier | 63(7.5) |
|  | Student | 190(22.6) |
|  | Working | 558(66.3) |
|  | Unemployed | 182(21.6) |
| Additional Chronic Comorbidity |  | 368(43.8) |
| Endometriosis Type | Superficial | 341(40.5) |
|  | Deep | 260(30.9) |
|  | Endometrioma | 121(14.4) |
|  | Neuropathic | 112(13.3) |
|  | Adenomyosis | 329(39.1) |
|  | Unknown | 188(22.4) |

Mean± Standard deviation; Number (%).

### Impact of War on Participants’ lives

82.8% of participants were directly affected by the war, suffering from at least one war-related stressor (median=2, IQR: 1-3) (**Supplementary Table 1**). 5% of participants were present at one of the attacked towns, at the “Nova” party, or at one of the military bases that were attacked on October 7, 2023. 24% suffered from persistent war-related stressors, being either evacuated from their home or had a close family member or friend murdered, injured or kidnapped during the war. 64.8% had second degree stressor through a friend or close relative affected by the war. 21.8% suffered economic damage. 48.9% were actively involved in war-related activities, either through military or civilian roles.

29% of participants reported impaired accessibility to healthcare services. Most accessibility issues were related to health system limitations (23.8%), including difficulty scheduling appointments, canceled imaging or surgery appointments, and limited access to complementary medicine. 15.9% of participants reported impaired accessibility due to military service, evacuation from their homes, and lack of personal availability (**Supplementary Table 2**). Participants reported worsening in different aspects of their daily activities since the beginning of the war, including eating habits (58.6%) and sleep patterns (71%), physical activity (44.9%), performing routine tasks (53.4%), participation in social gatherings (57%) and sharing their feelings concerning their disease (27.2%).

### Impact of War on Participants’ Mental Health

Participants’ mental health was profoundly affected, with a significant escalation in psychological distress following the war’s onset. 72.6% of participants experienced severe or very severe war-related stress (**Figure 1 A**). Moreover, depression rates nearly doubled, increasing from 39.6% before the war to 89.3% afterward, while anxiety levels rose from 34.7% to 94.1% (p<0.001 for both). Additionally, there was a significant change in the overall levels of depression & anxiety following the war. While prior to the war only 6.5% of participants had severe levels of depression & anxiety, at 2 months post-war beginning 71.1% of participants suffered from severe levels, gradually decreasing to 41.1% at ∼1 post-war begging (*p*<0.001) (**Figure 1 B**).

**Figure 1.**
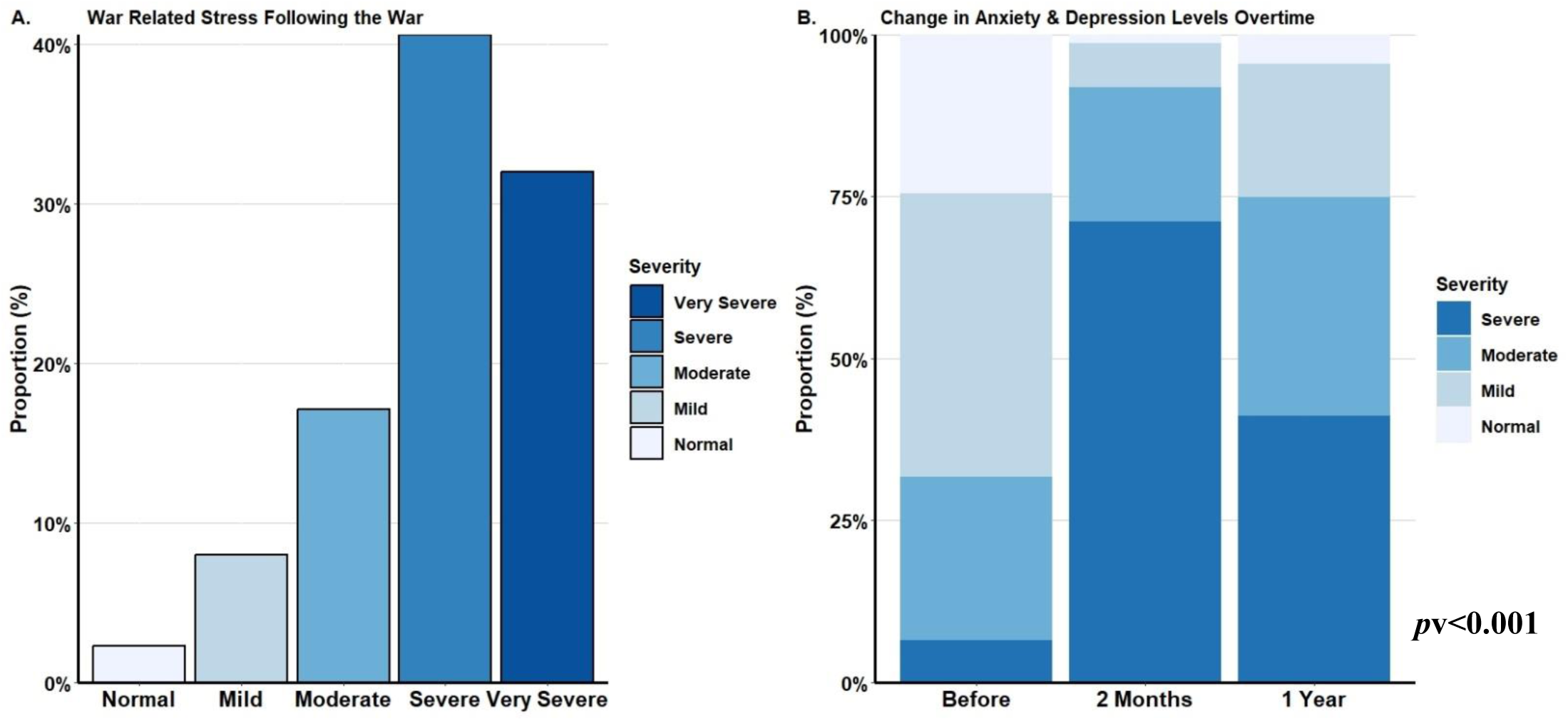
**(A.)** Severity of war related stress following the war and **(B.)** Severity of anxiety & depression following the war. Pv= p-value of Friedman test for paired repeated measures.

The war affected both participants’ positive and negative illness personification (**Table 2**). Both the overall negative IPS score, and all the specific negative personification items significantly deteriorated during the war. Participants had the lowest score two months post-war beginning, with a slight increase at ∼1 year though not returning to baseline. Interestingly, the overall positive IPS score, and most specific positive personification items were better at ∼1 year post-war beginning compared to baseline.

**Table 2.** The Effect of the War on the Perception of Endometriosis.

| Perception | Before | 2 Months | ~1 Year | p-value<sup>a</sup> |
| --- | --- | --- | --- | --- |
| **IPS Negative - Overall** | 2.8±1.1 | 3.0±1.2 | 2.9±1.1 | <0.001 |
| I feel like my disease is trying to destroy me | 2.9±1.4 | 3.2±1.4 | 3.1±1.5 | <0.001 |
| My disease is treacherous | 3.0±1.5 | 3.2±1.5 | 3.1±1.5 | <0.001 |
| My illness prevents me from becoming who I want to be | 2.9±1.5 | 3.1±1.5 | 3.1±1.5 | <0.001 |
| My illness shames me | 2.0±1.4 | 2.2±1.5 | 2.1±1.4 | <0.001 |
| My disease is malicious | 2.7±1.5 | 2.9±1.6 | 2.8±1.5 | <0.001 |
| I feel that my illness is like a formidable enemy | 3.2±1.5 | 3.4±1.5 | 3.3±1.4 | <0.001 |
| **IPS Positive - Overall** | 1.9±0.9 | 1.9±0.8 | 2.1±0.9 | <0.001 |
| My illness encourages me to grow | 2.4±1.4 | 2.1±1.3 | 2.5±1.4 | <0.001 |
| My illness is like a trusted friend | 1.4±1.0 | 1.4±1.0 | 1.5±1.0 | 0.023 |
| My illness reminds me of who I am | 2.3±1.4 | 2.3±1.5 | 2.5±1.5 | <0.001 |
| My illness connects me to myself | 2.3±1.4 | 2.3±1.4 | 2.6±1.5 | <0.001 |
| My illness teaches me like a wise old parent | 2.1±1.4 | 2.1±1.4 | 2.3±1.5 | <0.001 |
| My illness comforts me | 1.3±0.7 | 1.3±0.7 | 1.3±0.8 | 0.252 |

Mean± Standard deviation
The data were assessed by 5-point scale with 1 being "Highly Disagree" and 5 being "Highly Agree"
IPS = Illness Personification Scale
<sup>a</sup> Friedman Test for paired repeated measures.

Lastly, the number of war-related stressors were significantly correlated with participants’ war-related stress levels (r=0.24, *p*<0.001), and their anxiety & depression levels (r=0.09, *p*=0.034). Similarly, participants’ war-related stress levels were correlated with their anxiety & depression levels (r=0.36, *p*<0.001). Furthermore, participants’ negative perceptions of the disease (negative IPS levels) were correlated with both stress levels (r=0.23, *p*<0.001) and anxiety & depression levels (r=0.29, *p*<0.001).

### Impact of War on Participants’ Physical Health

The war also had a marked effect on participants’ physical health, particularly on endometriosis symptomatology. A total of 77.4% of participants reported a worsening in endometriosis symptoms, and 62.5% indicated a general decline in their overall medical condition during the conflict. Examination of various symptoms related to endometriosis revealed a significant deterioration in all symptoms following the war, including chronic pelvic pain, digestive and urinary symptoms, neuropathy, dyspareunia, menstrual pain, menstrual and intermenstrual bleeding, fatigue, and more (*p*<0.001 for all) (**Table 3**). Additionally, 38.9% and 14.4% of participants reported increased usage of pain and hormonal medications, respectively, indicating higher symptom management needs. Also, there was aggravation of participants’ medical condition over time, with initial escalation at two months post-war and a sustained but slightly reduced level a year later (*p*<0.001) (**Figure 2**). Worsening in participants’ medical condition was significantly correlated with the number of war-related stressors (r=0.16, *p*<0.001), their war-related stress levels (r=0.24, *p*<0.001), and their anxiety & depression levels (r=0.21, *p*<0.001).

**Figure 2.**
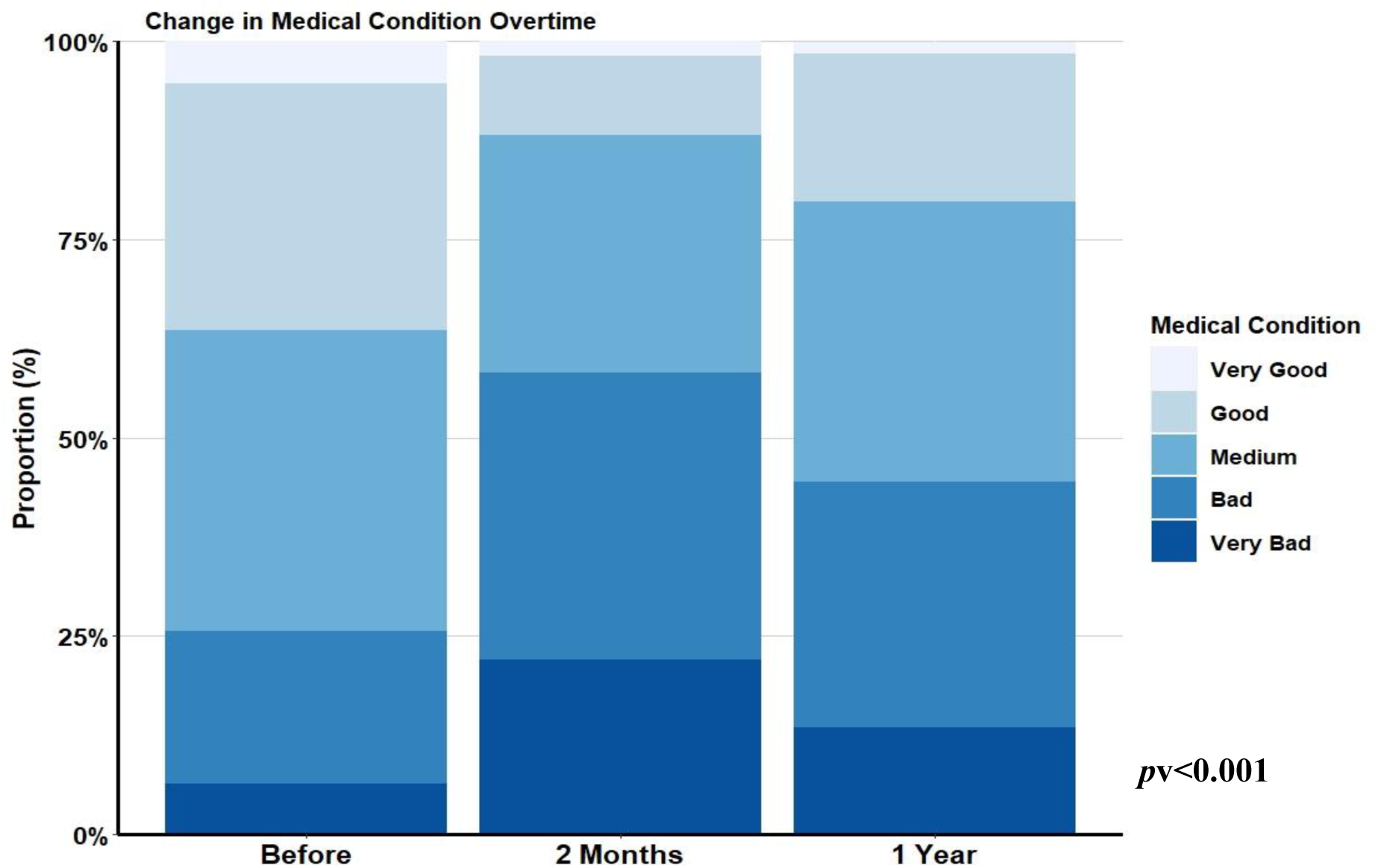
Change in medical condition overtime. Pv= p-value of Friedman test for paired repeated measures.

**Table 3.** The Effect of the War on Various Symptoms Related to Endometriosis.

| Symptom | Before | 2 Months | ~1 Year | p-value<sup>a</sup> |
| --- | --- | --- | --- | --- |
| Menstrual Pain | 3.7±1.0 | 4.2±1.0 | 4.0±1.0 | <0.001 |
| Chronic pelvic pain | 3.5±1.1 | 4.1±1.0 | 4.0±1.0 | <0.001 |
| Heavy menstrual bleeding | 3.6±1.2 | 3.9±1.2 | 3.8±1.2 | <0.001 |
| Intermenstrual bleeding | 2.7±1.4 | 3.5±1.3 | 3.2±1.4 | <0.001 |
| Changes in menstrual length | 2.9±1.3 | 3.6±1.2 | 3.6±1.3 | <0.001 |
| Digestive symptoms | 3.4±1.1 | 4.2±1.0 | 4.1±1.0 | <0.001 |
| Urinary system symptoms | 3.0±1.2 | 3.7±1.2 | 3.6±1.3 | <0.001 |
| Headache and/or migraines | 2.8±1.2 | 3.9±1.1 | 3.7±1.2 | <0.001 |
| Back and/or shoulder pain | 3.2±1.2 | 4.1±1.0 | 4.0±1.0 | <0.001 |
| Neuropathy | 3.1±1.2 | 3.8±1.2 | 3.7±1.3 | <0.001 |
| Vaginal Itching | 2.5±1.2 | 3.1±1.3 | 3.0±1.3 | <0.001 |
| Dyspareunia | 3.3±1.3 | 3.9±1.1 | 3.8±1.2 | <0.001 |
| Fatigue/exhaustion | 3.5±1.2 | 4.4±0.9 | 4.3±0.9 | <0.001 |
| Bloating | 3.4±1.2 | 4.0±1.1 | 4.0±1.1 | <0.001 |
| Sleep disorders | 3.0±1.3 | 4.3±0.9 | 4.0±1.0 | <0.001 |

Mean± Standard deviation
Symptoms severity was assessed on a 5-point scale, with 1 being the best and 5 being the worst.
<sup>a</sup> Friedman Test for paired repeated measures.

Finally, we used multivariable logistic regression to examine the impact of war-related stressors, participants’ mental health statue, and impaired healthcare accessibility on participants’ physical health after adjusting to potential confounders (**Figure 3**). The same trend was observed in both endometriosis symptoms (ES) and medical condition (MC). In both, the number of war-related stressors, war-related stress level, depression & anxiety level, impaired healthcare accessibility and negative illness personification were significantly associated with worse outcomes (<u>War-Related Stressors</u>: aOR_ES_=1.24, 95CI=1.07-1.45; aOR_MC_=1.20, 95CI=1.04-1.39; <u>War-Related Stress</u>: aOR_ES_=1.83, 95CI=1.50-2.22; aOR_MC_=1.84, 95CI=1.51-2.23; <u>Depression & Anxiety</u>: aOR_ES_=2.00, 95CI=1.52-2.63; aOR_MC_=2.06, 95CI=1.57-2.71; <u>Impaired Healthcare Accessibility</u>: aOR_ES_=1.64, 95CI=1.04-2.57; aOR_MC_=1.81, 95CI=1.16-2.84; <u>Negative IPS</u>: aOR_ES_=1.37, 95CI=1.12-1.67; aOR_MC_=1.48, 95CI=1.21-1.81). In contrast, positive illness personification showed protective effects (aOR_ES_=0.77, 95CI=0.59-0.99; aOR_MC_=0.81, 95CI=0.63-1.04).

**Figure 3.**
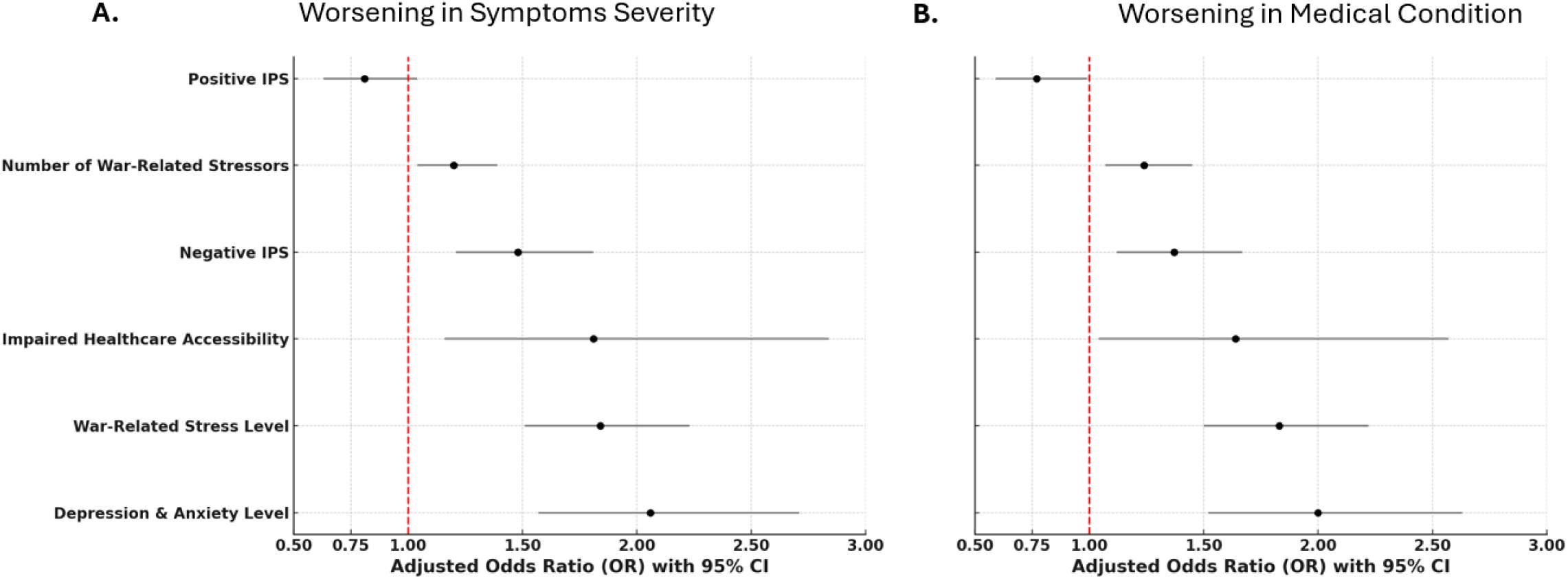
The Impact of War on Worsening Endometriosis Symptoms Severity & Medical Condition. (**A.)** Worsening in symptoms severity - multivariable logistic regression adjusted to age, army service, education, healthcare accessibility, new endometriosis finding, new medical diagnosis, pregnancy, fertility treatment, loss of a relative or close friend. (**B.)** Worsening in medical condition - multivariable logistic regression adjusted to age, army service, religious, healthcare accessibility, new endometriosis finding, new medical diagnosis, surgery, loss of a relative or close friend. CI = Confidence Interval

## Discussion

This large-scale population-based study highlights the profound impact of war and war-related stress on both the physical and mental health of endometriosis patients. The results of this study demonstrate that patients’ lives were deeply affected by the war, with most encountering direct exposure to war-related stressors, impaired access to healthcare services, and significant disruptions to daily routines. Moreover, mental health was severely affected, with most experiencing high levels of war-related stress and a dramatic increase in anxiety and depression rates compared to pre-war levels. The war also led to significant deterioration in patients’ illness perception. Finally, physical health deteriorated substantially, with most reporting worsening of endometriosis symptoms and overall medical decline. Notably, the deterioration in both physical symptoms and overall medical condition showed significant associations with both psychological distress levels and the number of war-related stressors, even after adjusting for potential confounders such as impaired healthcare accessibility.

The war had a substantial impact on participants’ mental health, manifested by elevated levels of war-related stress, significant increases in anxiety and depression rates, and deterioration in illness perception. A dose-dependent correlation was observed, with higher exposure to war-related stressors correlating with greater psychological distress, including more severe anxiety, depression, and stress levels. These findings align with previous research demonstrating the substantial impact of war on mental health.^24–29^ Recent studies conduct in Israel following the war demonstrated increased rates of PTSD, anxiety, depression, and psychological distress,^36–39^ particularly among women and individuals with greater exposure to traumatic events.^29,36,37,48^ In addition, these studies found that traumatic events had a more pronounced impact on individuals with pre-existing psychological difficulties,^36^ further emphasizing the vulnerability of women with endometriosis, who already face an elevated risk of mental health disorders and higher levels of perceived stress compared to the general population.^4–12,40^ Our study showed that while patients suffered from a dramatic increase in depression and anxiety following the war, it gradually decreased at ∼1 year post-war beginning, though still remaining significantly higher than before its start. This finding aligns with a previous meta-analysis demonstrating a downward trend after a given war had ended.^49^ Interestingly, participants’ positive illness personification was better at ∼1 year post-war beginning compared to baseline. Previous studies found that positive IPS is positively correlated with both dissociation and resilience. Indeed, dissociation may occur in response to traumatic events such as war, and may explain the increase in positive IPS following the war.^45^ Nevertheless, this finding may also be explained by posttraumatic growth and increase in participants’ resilience due to the war, as found in a recent study demonstrating a national rise in resilience and collective post-traumatic growth following the October 7 war.^50^

Alongside the impact on participants’ mental health, the war also significantly affected participants’ physical well-being, with the vast majority reporting a marked deterioration in their overall medical condition and endometriosis symptoms. Moreover, a considerable number of participants reported increased reliance on pain and hormonal medications, reflecting heightened symptom management needs. Notably, the deterioration in both endometriosis symptoms and overall medical condition showed significant associations with both psychological distress levels and the number of war-related stressors, even after adjusting for potential confounders including healthcare accessibility, suggesting that external stressors may affect and exacerbate symptoms of chronic medical conditions such as endometriosis. This finding is in line with previous studies demonstrating the effect of extreme stressful experiences such a the COVID pandemic and war on the physical and mental health of patients with chronic disease such as endometriosis and multiple sclerosis.^17,40,51,52^ Stress has been shown to disrupt homeostasis by modulating hormonal pathways, immune responses, and inflammatory mechanisms, all of which play crucial roles in the pathophysiology of endometriosis, leading to increase in the size and severity of the endometrial lesions and exacerbation of inflammatory parameters.^13–17,19,20^ Moreover, chronic stress can increase nerve fiber density, amplifying pain perception and inflammatory processes in endometriosis patients.^20–22^ In contrast, stress reduction has been linked to decreased lesion size emphasizing the critical role of stress in modulating disease progression.^22,23^ These findings emphasize the need for early identification of high-risk patients and implementation of comprehensive support strategies that address both symptom management and psychological well-being during times of crisis

Our research has several limitations. First, the cross-sectional nature of the survey limits our ability to establish causality between war-related stress to mental health deterioration and symptom exacerbation. Second, while our study had a large sample size, participants were recruited primarily through social media platforms and support groups, which may have led to selection bias towards more engaged patients or those experiencing more severe symptoms. Additionally, the reliance on self-reported data and retrospective assessment of pre-war symptoms may introduce recall bias. The study’s timing during an ongoing conflict might have influenced participants’ responses, particularly regarding psychological distress. Furthermore, while we attempted to adjust for various confounding factors, other factors not assessed in the current study may have also affected symptom severity.

## Conclusions

In conclusion, this study demonstrates the profound impact of war on both the mental and physical health of endometriosis patients. Our findings underscore the importance of maintaining healthcare accessibility and implementing targeted support strategies that address both the physical and psychological well-being of this vulnerable population during national crises

## Data Availability

All data produced in the present study are available upon reasonable request to the authors

