## Supplementary material for "Silent Battles: The Impact of War on the Mental and Physical Health of Endometriosis Patients": Supplemntary Tables 1-4

**Supplementary Table 1. Exposure to War-Related Stressful Events**

| Variable | Number (%) |
|---|---|
| **Direct Stressor** | **42(5.0)** |
| Were you present at one of the attacked settlements, at the Nova party, or at one of the military bases that were attacked during the October 7 attack? | 42(5.0) |
| **Persistent Stressor** | **202(24.0)** |
| Where you evacuated from you home since the 07.10 attack? | 45(5.4) |
| Was one of your close family members or a close friend murdered, injured or kidnapped in the October 7 attack, or throughout the war? | 175(20.8) |
| **Secondary Stressor** | **545(64.8)** |
| Have you been directly exposed to the experiences of a person affected by the October 7 attack (whether as part of providing a therapeutic response or as part of a personal relationship)? | 359(42.7) |
| Has your spouse or first degree relative served in the reserves since October 7? | 356(42.3) |
| **Economic Stressor** | **183(21.8)** |
| Was your home/property damaged during the war? | 23(2.7) |
| Did you experience significant economic damage following the war? | 169(20.1) |
| **Significant Action** | **411(48.9)** |
| Have you served in reserves since October 7? | 75(8.9) |
| Did you take an active part in promoting tasks related to the war? | 376(44.7) |

**Supplementary Table 2. Impaired Accessibility to Health Services Due to the War**

| Constraint Type | Number (%) |
|---|---|
| **Overall** | **188(29.0)** |
| **Health System Constraints** | **154(23.8)** |
| Difficulty finding an appointment for imaging | 84(13.0) |
| Imaging appointment cancelled | 27(4.2) |
| Surgery Appointment Postponed | 13(2.0) |
| I did not receive a response to a request for a prescription from a specialist doctor to treat the disease | 58(9.0) |
| I had difficulty finding appointments for follow-up and/or diagnosis at a specialist clinic | 127(19.6) |
| My appointment for follow-up and/or diagnosis at a specialist clinic was cancelled | 65(10.0) |
| I had difficulty finding appointments for complementary medicine | 85(13.1) |
| My appointment for complementary medicine was Cancelled | 39(6.0) |
| **Patient Constraints** | **103(15.9)** |
| I requested to cancel appointments in light of the situation and/or due to military service | 88(13.6) |
| I requested to cancel appointments due to evacuation from my home | 17(2.6) |
| I requested to cancel appointments due to a lack of availability | 68(10.5) |

**Supplementary Table 3. Association between Participant Characteristics and Endometriosis Symptoms Severity – Univariate Analysis**

| Variable | | Odds Ratio (OR) | 95% CI | p-value[a] |
|---|---|---|---|---|
| Age | | 0.98 | 0.96-0.99 | **0.031** |
| Jewish Ethnicity | | 0.79 | 0.32-1.94 | 0.600 |
| Religious | | 0.76 | 0.54-1.06 | 0.108 |
| Children | Children >18 | 0.63 | 0.29-1.35 | 0.234 |
| | Children<18 | 0.76 | 0.52-1.12 | 0.167 |
| Occupation | Soldier/Reserves/Service Year | 2.08 | 1.01-4.28 | **0.046** |
| | University Student | 1.48 | 0.97-2.27 | 0.069 |
| | Working | 0.89 | 0.62-1.27 | 0.523 |
| Higher Education | | 0.64 | 0.46-0.90 | **0.010** |
| Additional Chronic Comorbidity | | 1.21 | 0.86-1.69 | 0.267 |
| Endometriosis Type | Superficial | 1.16 | 0.83-1.62 | 0.400 |
| | Deep | 1.09 | 0.76-1.56 | 0.633 |
| | Endometrioma | 0.86 | 0.54-1.34 | 0.487 |
| | Neuropathic | 1.62 | 0.95-2.75 | 0.079 |
| | Adenomyosis | 0.84 | 0.60-1.17 | 0.301 |
| | Unknown | 1.02 | 0.69-1.50 | 0.935 |
| Impaired Accessibility to Health Services | | 1.80 | 1.17-2.77 | **0.008** |
| New Diagnosis Since War | New Endometriosis Finding | 2.51 | 1.60-3.93 | **<0.001** |
| | New Medical Diagnosis | 1.82 | 1.14-2.91 | **0.013** |
| War-Related Stressors | Direct Stressor | 0.94 | 0.45-2.08 | 0.969 |
| | Persistent Stressor | 1.60 | 1.05-2.45 | **0.029** |
| | Secondary Stressor | 1.42 | 1.01-1.99 | **0.045** |
| | Economic Stressor | 2.04 | 1.29-3.23 | **0.002** |
| | Significant Action | 1.39 | 0.99-1.94 | 0.056 |
| | Number of Stressors | 1.30 | 1.15-1.48 | **<0.001** |
| IPS | IPS Negative | 1.45 | 1.20-1.74 | **<0.001** |
| | IPS Positive | 0.80 | 0.63-1.02 | 0.073 |
| War-Related Stress Level | | 1.76 | 1.49-2.07 | **<0.001** |
| Fear & Anxiety Level | | 1.14 | 1.06-1.22 | **<0.001** |
| | Divorce / Breakup | 1.43 | 0.58-3.50 | 0.435 |
| | Marriage | 0.87 | 0.34-2.23 | 0.771 |
| | Surgery | 0.95 | 0.57-1.60 | 0.851 |
| | Pregnancy | 0.38 | 0.19-0.74 | **0.004** |
| | Vaginal/Caesarean Delivery | 0.49 | 0.19-1.26 | 0.139 |
| | Fertility Treatments | 0.37 | 0.22-0.63 | **<0.001** |
| | Layoffs or job changes | 1.11 | 0.74-1.65 | 0.615 |
| | Home change not due to the war | 1.12 | 0.63-2.00 | 0.695 |
| | Loss of a relative or close friend not due to the war | 2.71 | 1.22-6.03 | **0.015** |

[a] Univariate logistic regression

OR = Odds Ratio, CI = Confidence Interval

**Supplementary Table 4. Association between Participant Characteristics and Worsening of Medical Condition – Univariate Analysis**

| **Variable** | | **Odds Ratio (OR)** | **95% CI** | **p-value[a]** |
|---|---|---|---|---|
| Age | | 0.99 | 0.97-1.01 | 0.167 |
| Jewish Ethnicity | | 0.99 | 0.48-2.08 | 0.998 |
| Religious | | 0.65 | 0.48-0.88 | **0.005** |
| Children | Children >18 | 0.89 | 0.63-1.26 | 0.519 |
| | Children<18 | 0.95 | 0.45-1.98 | 0.889 |
| Occupation | Soldier/Reserves/Service Year | 1.68 | 0.97-2.91 | 0.063 |
| | University Student | 1.40 | 0.98-1.99 | 0.065 |
| | Working | 0.91 | 0.67-1.24 | 0.551 |
| Higher Education | | 1.24 | 0.93-1.66 | 0.147 |
| Additional Chronic Comorbidity | | 0.82 | 0.62-1.10 | 0.190 |
| Endometriosis Type | Superficial | 1.14 | 0.85-1.52 | 0.397 |
| | Deep | 0.76 | 0.56-1.04 | 0.087 |
| | Endometrioma | 0.82 | 0.55-1.22 | 0.329 |
| | Neuropathic | 0.87 | 0.58-1.32 | 0.508 |
| | Adenomyosis | 0.87 | 0.65-1.17 | 0.361 |
| | Unknown | 1.23 | 0.87-1.74 | 0.246 |
| Impaired Accessibility to Health Services | | 1.16 | 0.81-1.66 | 0.413 |
| New Diagnosis Since War | New Endometriosis Finding | 1.34 | 0.96-1.88 | 0.087 |
| | New Medical Diagnosis | 1.57 | 1.07-2.31 | **0.022** |
| War-Related Stressors | Direct Stressor | 1.11 | 0.56-2.22 | 0.759 |
| | Persistent Stressor | 1.54 | 1.08-2.19 | **0.017** |
| | Secondary Stressor | 1.15 | 0.85-1.55 | 0.375 |
| | Economic Stressor | 1.78 | 1.23-2.57 | **0.002** |
| | Significant Action | 1.37 | 1.02-1.83 | **0.035** |
| | Number of Stressors | 1.24 | 1.11-1.38 | **<0.001** |
| IPS | IPS Negative | 1.07 | 0.91-1.24 | 0.413 |
| | IPS Positive | 0.88 | 0.71-1.09 | 0.254 |
| War-Related Stress Level | | 1.48 | 1.27-1.71 | **<0.001** |
| Fear & Anxiety Level | | 1.24 | 1.16-1.34 | **<0.001** |
| | Divorce / Breakup | 1.16 | 0.57-2.36 | 0.687 |
| | Marriage | 1.73 | 0.67-4.43 | 0.256 |
| | Surgery | 0.61 | 0.39-0.94 | **0.026** |
| | Pregnancy | 0.83 | 0.42-1.64 | 0.599 |
| | Vaginal/Caesarean Delivery | 0.53 | 0.21-1.32 | 0.175 |
| | Fertility Treatments | 0.66 | 0.39-1.11 | 0.119 |
| | Layoffs or job changes | 1.11 | 0.78-2.56 | 0.567 |
| | Home change not due to the war | 0.83 | 0.51-1.34 | 0.441 |
| | Loss of a relative or close friend not due to the war | 1.71 | 0.97-2.99 | 0.062 |

[a] Univariate logistic regression

OR = Odds Ratio, CI = Confidence Interval
